# Can a rapid-access vascular limb salvage service improve one-year amputation outcomes for patients with chronic limb-threatening ischaemia?

**DOI:** 10.1101/19013037

**Authors:** A.T.O. Nickinson, J. Dimitrova, J.S.M. Houghton, L. Rate, S. Dubkova, H. Lines, L.J. Gray, S. Nduwayo, T. Payne, R.D. Sayers, R.S.M. Davies

**Author notes:** Corresponding author: Mr A. Nickinson (, @AndrewNickinson), Department of Cardiovascular Sciences, University of Leicester, Glenfield Hospital, Groby Road, LE3 9QP, UK, Original study.

## Abstract

**Background:** Vascular limb salvage services can potentially improve outcomes for patients with chronic limb-threatening ischaemia (CLTI), although their description within the literature is limited. This study aims to evaluate the 12-month outcomes for an outpatient-based vascular limb salvage (VaLS) clinic and investigate times-to-treatment.

**Methods:** An analysis of a prospectively maintained database, involving all patients diagnosed with CLTI within the VaLS clinic from February 2018-February 2019, was undertaken. Data were compared to two comparator cohorts, identified from coding data; 1) patients managed prior to the clinic, between May 2017-February 2018 (Pre-Clinic’ [PC]), and 2) patients managed outside of clinic, between February 2018-February 2019 (‘Alternative Pathways’ [AP]). Freedom from major amputation at 12 months was the primary outcome. Kaplan-Meier plots and adjusted Cox’s proportional hazard models (aHR) were utilised to compare outcomes.

**Results:** Five-hundred and sixty-six patients (VaLS=158, AP=173, PC=235) were included (median age=74 years). Patients managed within the VaLS cohort were significantly more likely to be free from major amputation (90.5%) compared to both the AP (82.1%, aHR 0.52, 95% CI 0.28-0.98, p=.041) and the PC (80.0%; aHR 0.50, 95% CI 0.28-0.91, p=.022) cohorts at 12 months, after adjustment for age, disease severity and presence of diabetes.

**Conclusions:** A limb salvage clinic may help improve the rate of major amputation and provides a reproducible model which delivers timely vascular assessment in an ambulatory setting. Further evaluation is required to assess longer-term outcomes.

## Introduction

Despite efforts to improve the care given to patients with chronic limb-threatening ischaemia (CLTI), comparatively little attention has been paid to evolving the clinical pathways through which patients are managed. The focus on multi-disciplinary team (MDT) working remains one of the most beneficial changes in this area, particularly for patients with neuro-ischaemic diabetic foot ulcers (DFU).^1,2^ Even in the age of MDT working, lengthy time delays in the management of CLTI occur, with these delays having a detrimental effect on patient outcomes.^3^ Reasons cited for these delays include difficulties accessing specialist vascular services by ‘community’ and ‘non-specialist’ healthcare professionals, and unnecessary assessment by ‘non-specialist’ professionals prior to referral.^4-6^

In an attempt to help improve care and reduce rates of major amputation, the Vascular Society of Great Britain and Ireland (VSGBI) recently published its ‘Peripheral Arterial Disease Quality Improvement Framework’ (PAD QIF), which for the first time stipulates target times for the assessment and revascularisation of patients with CLTI (Figure 1).^7^ Whilst the ambition of these targets is laudable, they are by the VSGBI’s own description “deliberately challenging” and currently it remains unclear how hospitals will achieve compliance.^7^

**Figure 1.**
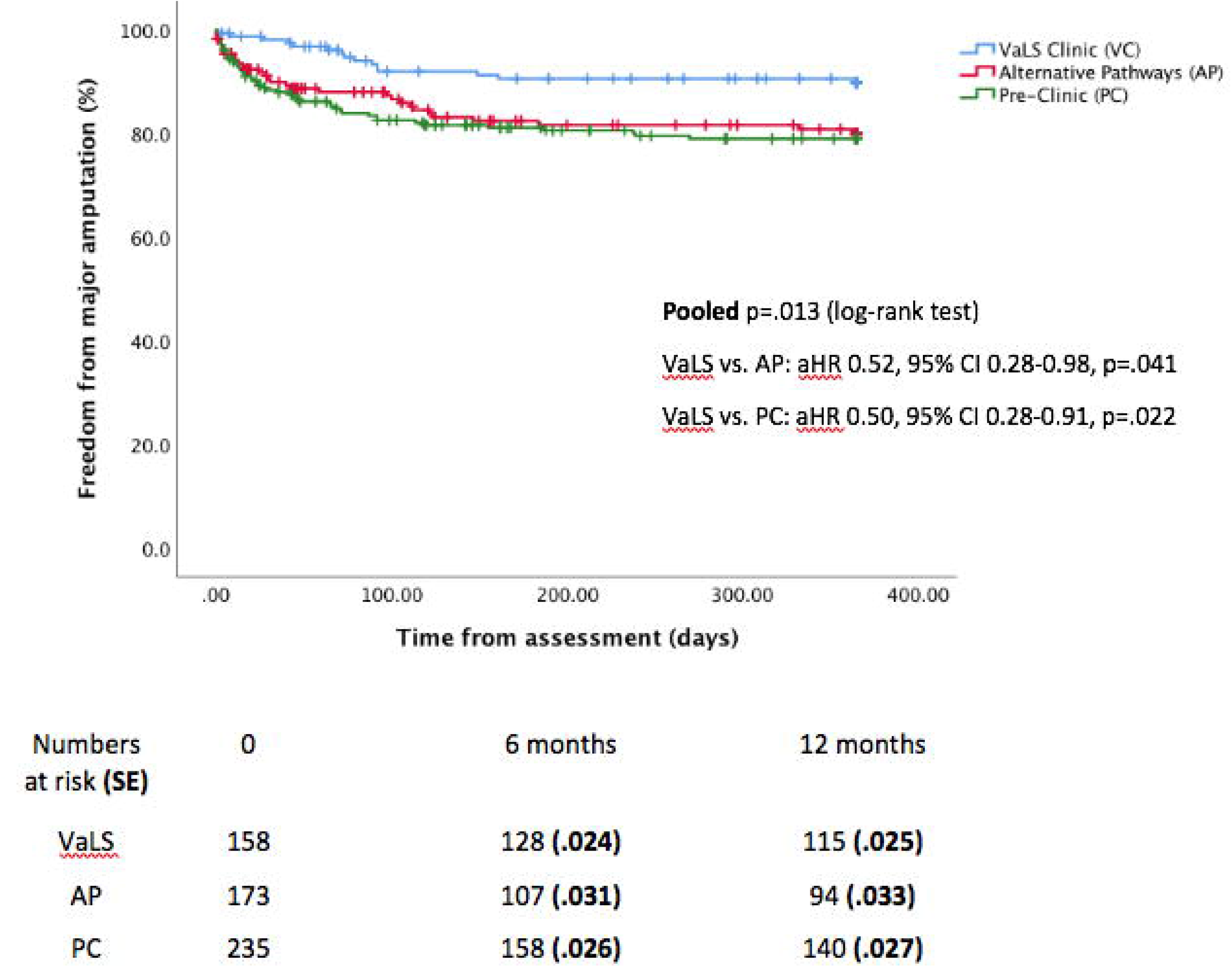
– Vascular Society of Great Britain and Ireland’s time-to-treatment targets. *‘Admitted Patient’ and ‘Non-Admitted Patient’ pathways within the Vascular Society of Great Britain and Ireland’s ‘A best practice clinical care pathway for peripheral arterial disease ‘ (London, 2019*).^7^ *Reproduced with kind permission*.

Specialist vascular limb salvage services are one model of care which potentially provide a solution to these challenges. Although services can take different forms (reflecting the diverse nature of healthcare systems), at their core they provide rapid-access to specialist vascular assessment and revascularisation of an ischaemic limb, along with debridement of necrosis and drainage of infection.^8-10^ In February 2018, the Leicester Vascular Institute in the United Kingdom opened a vascular limb salvage (VaLS) clinic to help improve amputation outcomes for patients with CLTI.

To date, not only are examples of these services within the literature limited, but many have key methodological weakness, which affects the validity of their conclusion.^11^ Furthermore, no studies have investigated the effect of limb salvage services on times-to-treatment. The aims of this study are therefore to, a) describe the service provided by the VaLS clinic, b) evaluate the 12-months limb salvage outcomes for patients managed within the clinic’s first year of operation and c) investigate the effect of limb salvage services on times-to-treatment.

## Methodology

### Setting

Prior to the inception of the VaLS clinic, patients with CLTI were managed through the traditional in/outpatient model of care. Outpatient pathways facilitated the initial management of patients with rest pain and/or dry necrosis, requiring formal written referrals from community/non-specialist healthcare professionals. In cases of suppurative infection, wet gangrene or sepsis, patients were admitted directly to the Leicester Vascular Institute as an emergency to facilitate treatment. All patients had access to lower limb arterial duplex ultrasonography (+/- computerised tomography angiography [CTA]), with subsequent decisions on treatment discussed in weekly MDT meetings, where necessary.

The VaLS clinic opened on 7^th^ February 2018 with the purpose of providing a rapid, open-access, ‘one-stop’, outpatient-based vascular limb salvage clinic for adult patients with suspected CLTI. The primary aim of the clinic was to reduce time from referral to revascularisation (≤14 days) and reduce rates of major amputation.

The clinic is led by two vascular specialist nurses who manage referrals and complete initial patient assessment. A dedicated vascular scientist works within the clinic to perform lower limb arterial duplex ultrasonography. Assessments are available for up to four patients per day, Monday-Friday. The clinic also has access to reserved, fast-track CTA (same-day) and rapid-access to angio-suite and hybrid theatre appointments for revascularisation.

Referral to the clinic is made via telephone, email or online referral portal from any healthcare professional managing a patient. The clinic offers an ‘open access’ policy and as such clinical suspicion of CLTI is the only referral criterion. The clinic was designed to cover the Leicestershire and Rutland region of the United Kingdom (approximately 1.1 million individuals), however referrals from outside this area are also accepted.

On initial consultation patients undergo vascular assessment, formulated in line with contemporary guidelines.^10^ Suitable patients undergo ankle-brachial pressure index measurement, with toe-brachial pressure index undertaken if incompressible or unreliable. Arterial duplex ultrasonography is subsequently performed, imaging from the common femoral artery (including aorto-iliac inflow, if indicated) to pedal arch outflow. A Society of Vascular Surgery WIfI (wound, ischaemia, foot infection) score is also recorded where possible and converted into stages to define the risk of major amputation and benefit of revascularisation (stage 1=very low risk, 2=low risk, 3=moderate risk, 4=high risk).^12^

Following initial assessment all patients are reviewed in the same consultation by a consultant vascular surgeon. Decisions regarding revascularisation are taken with the aim of establishing in-line flow to the affected limb. The choice of endovascular or open-first strategy is based upon the assessment of anatomy and suitability for endovascular or open revascularisation including suitability of a conduit vein, patient’s functional status and perioperative risk (based upon the surgeon’s subjective assessment), and patient preference. All decisions requiring endovascular intervention are made in conjunction with local endovascular specialists. When required, patients undergo expedited, outpatient pre-operative assessment, including echocardiogram and pulmonary function testing, to help objectively define perioperative risk.

Endovascular procedures are predominantly undertaken under local anaesthetic on an expedited outpatient basis, with patients requiring open procedures also able to receive a planned operation date at the time of consultation. In situations where urgent treatment is required (e.g. drainage of suppurative infection), patients are admitted directly to the Leicester Vascular Institute.

All clinical decisions are discussed in a daily team meeting involving vascular surgeons and nurse specialists, and allied health professionals. In cases of complex revascularisation or management decisions, patients are discussed in a weekly MDT meeting (involving interventional radiologists, anaesthetists, and if required, specialists from diabetology and internal medicine).

Where appropriate, secondary preventative therapies (i.e. antiplatelet and lipid lowering agents) are commenced. If no revascularisation or debridement is required, or in cases where patients do not reach the diagnostic threshold for CLTI (e.g. claudication), patients are discharged back to the referring clinician or routine follow-up in vascular outpatient clinic is arranged.

Given the operational hours of the clinic, the emergency admission pathway is available for emergency out-of-hours referrals when required.

### Study design and population

A retrospective cohort study, reviewing a prospectively maintained clinic database was undertaken. All consecutive patients diagnosed with CLTI within the VaLS clinic between 7^th^ February 2018 (inception date) and 6^th^ February 2019 were included. Patients were defined as having a diagnosis of CLTI based upon clinical history (tissue loss or rest/night pain for ≥2 weeks) in conjunction with objective evidence of PAD (equivalent of WIfI ischaemia grade ≥1). These patients formed the ‘VaLS Clinic’ (VaLS) cohort.

To provide context, patients were compared to two comparator cohorts. Firstly, all patients managed for in the 9-month period prior to the inception of the clinic (1^st^ May 2017 - 6^th^ February 2018), termed the ‘Pre-Clinic’ (PC) cohort. This period was chosen to give the most comparable cohort prior to opening of the clinic but is after a major relocation of the Leicester Vascular Institute, which finished in April 2017. Secondly, all patients managed after inception of the clinic (7^th^ February 2018 - 6^th^ February 2019) but via alternative care pathways and not first through the VaLS clinic (i.e. elective outpatient clinics, or at times when the clinic is not open, such as weekends). This cohort was termed the ‘Alternative Pathways’ (AP) cohort.

Patients for both comparator cohorts were identified from searching Hospital Episode Statistics (HES) data using combinations of ICD-10 (I70.20 and I70.21; atherosclerosis of lower limbs with/without gangrene) and OPCS-4 (X09.-X12.1; major/minor lower limb amputation) codes. Data were cross-referenced with electronic patient records and a local, prospectively maintained vascular registry database to ensure accuracy. Patients diagnosed with acute limb ischaemia, intermittent claudication, venous disease, DFU without steno-occlusive PAD, or non-atherosclerotic PAD were excluded.

### Baseline data and outcomes

Baseline demographics including age, sex and comorbidities documented at date of assessment (hypertension, ischaemic heart disease, prior stroke/transient ischaemic attack, diabetes mellitus) were collected from patient electronic records. Where available, the use of best medical therapy at or close to the date of assessment (anti-platelet or anti-coagulation and lipid lowering medications) were also collected. Given that patients within the comparator cohorts were identified through coding and electronic records, it was not possible to ascertain individual WIfI scores. Therefore, disease severity for all patients at assessment was graded using Rutherford scores.

Referral dates and subsequent procedural data (first attempted revascularisation, minor and major amputation) and mortality were identified. A major amputation was defined as an amputation proximal to the ankle joint. All patients were followed-up until 12 months post assessment or death (whichever occurred first).

Freedom from major amputation at 12 months was defined as the primary outcome of the study. Secondary outcomes were amputation free-survival at 12 months and times-to-treatment (assessment to first attempted revascularisation). While referral dates are collected prospectively within the VaLS clinic, this information is not accurately available in retrospect and therefore this was not assessed within the two comparative cohorts.

### Statistical analysis

Continuous baseline variables and times-to-treatment were examined for normality using histogram plots and presented as either means (with standard deviation [SD]) or medians (with interquartile ranges [IQR]). Categorical variables are presented as frequencies (with percentages). Comparison between cohorts was undertaken using Pearson’s chi-square, Student’s-t or Mann-Whitney U-tests, as appropriate. All analyses were undertaken on a ‘per patient’ basis, as such patients who underwent multiple assessments were analysed based upon their first clinical assessment, and in the cohort in which this assessment occurred. Outcomes of major amputation were analysed irrespective the side of amputation.

Kaplan-Meier curves were plotted and cohorts compared using the log-rank rest. Cox’s proportional hazard models were also utilised to calculate hazard ratios (HR) with 95% confidence intervals (CIs). The baseline characteristics of age, diabetes, use of anti-platelet and lipid lowering therapy, and disease severity (Rutherford score) were selected *a priori* to be added into a final adjusted (aHR) model, given their potentials effect on amputation outcomes.^13^ All statistical analysis was performed using SPSS v26.0 (IBM Corp., Armonk, USA). P-values ≤.05 were deemed statistically significant.

## Results

Six-hundred and eighteen consecutive patients managed for CLTI were identified across all three cohorts (VaLS=180, AP=203, PC=235 patients). Of these, 52 patients underwent assessment in multiple cohorts (VaLS=22, AP=30 patients). Taking account of the cohort in which patients underwent their first assessment, analysis of outcomes was performed on 158 patients within the VaLS, 173 patients within the AP and 235 patients within the PC cohorts (total=566 patients).

Comparison of baseline characteristics is shown in Table 1. Overall there was good comparability between the cohorts, although of note a lower proportion of patients within the VaLS cohorts were receiving best medical therapy (principally lipid lowering therapy) at assessment. In all three cohorts, the vast majority of patients presented with tissue loss in the form of minor ulceration (Rutherford stage 5) and no statistically significant difference in disease severity was observed between cohorts.

**Table 1.**
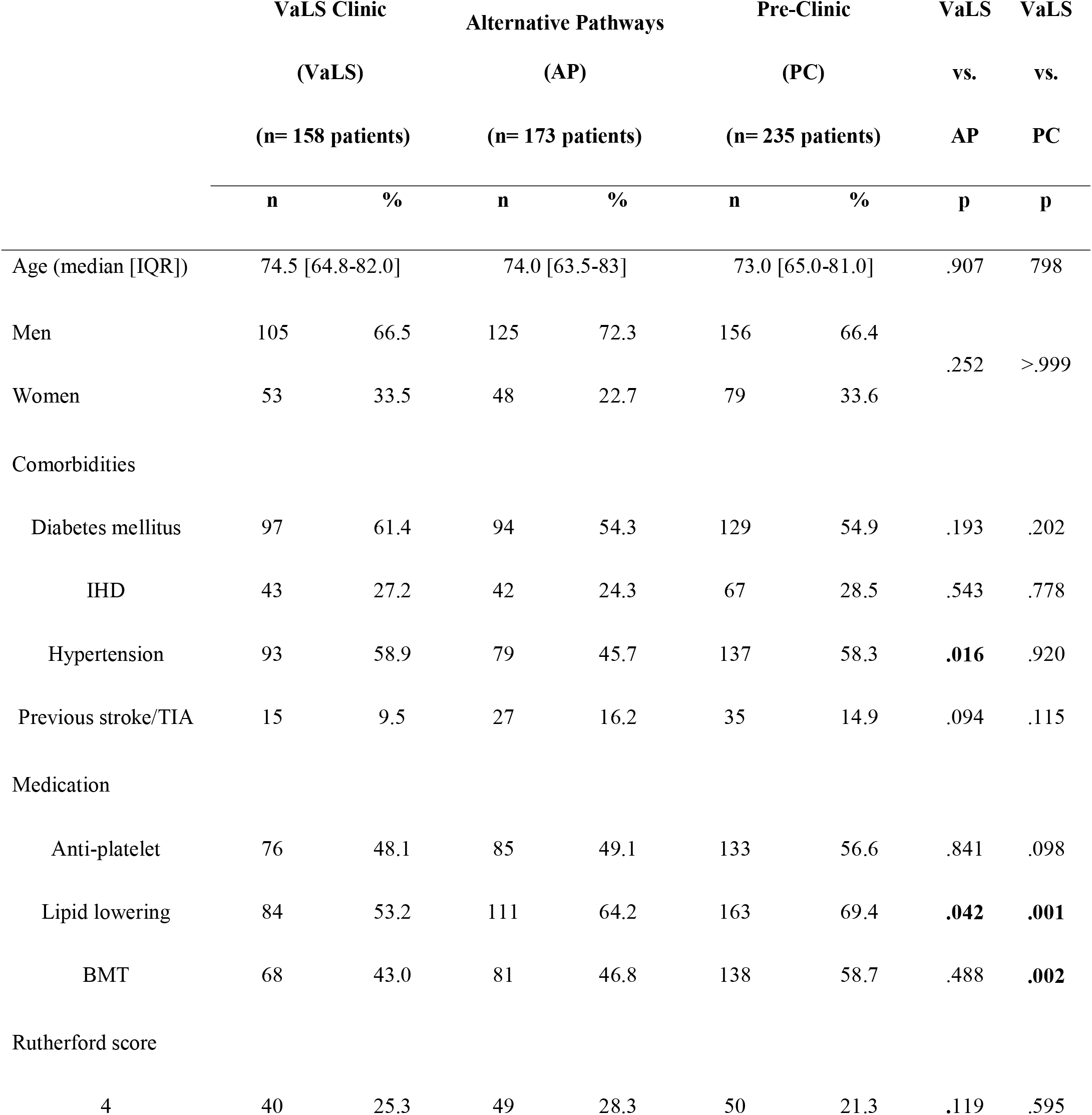
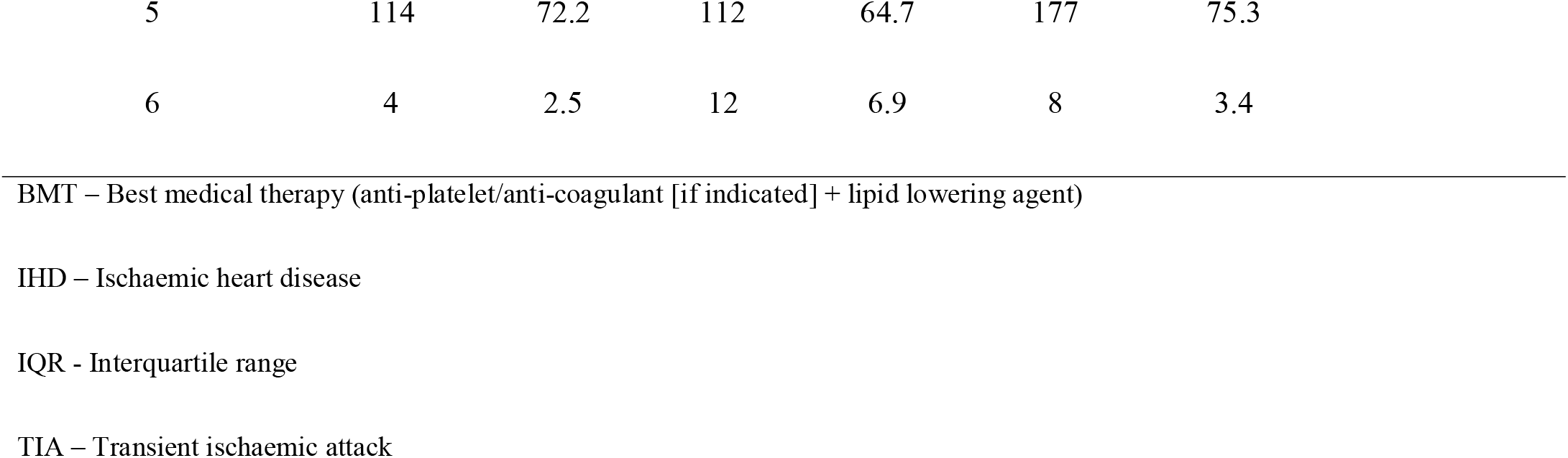
–Baseline characteristics of study cohorts.

|  | VaLS Clinic |  | Alternative Pathways |  | Pre-Clinic |  | VaLS | VaLS |
| --- | --- | --- | --- | --- | --- | --- | --- | --- |
|  | (VaLS) |  | (AP) |  | (PC) |  | vs. | vs. |
|  | (n= 158 patients) |  | (n= 173 patients) |  | (n= 235 patients) |  | AP | PC |
|  | n | % | n | % | n | % | p | p |
| Age (median [IQR]) | 74.5 [64.8-82.0] |  | 74.0 [63.5-83] |  | 73.0 [65.0-81.0] |  | .907 | .798 |
| Men | 105 | 66.5 | 125 | 72.3 | 156 | 66.4 | .252 | >.999 |
| Women | 53 | 33.5 | 48 | 22.7 | 79 | 33.6 |  |  |
| Comorbidities |  |  |  |  |  |  |  |  |
| Diabetes mellitus | 97 | 61.4 | 94 | 54.3 | 129 | 54.9 | .193 | .202 |
| IHD | 43 | 27.2 | 42 | 24.3 | 67 | 28.5 | .543 | .778 |
| Hypertension | 93 | 58.9 | 79 | 45.7 | 137 | 58.3 | **.016** | .920 |
| Previous stroke/TIA | 15 | 9.5 | 27 | 16.2 | 35 | 14.9 | .094 | .115 |
| Medication |  |  |  |  |  |  |  |  |
| Anti-platelet | 76 | 48.1 | 85 | 49.1 | 133 | 56.6 | .841 | .098 |
| Lipid lowering | 84 | 53.2 | 111 | 64.2 | 163 | 69.4 | **.042** | **.001** |
| BMT | 68 | 43.0 | 81 | 46.8 | 138 | 58.7 | .488 | **.002** |
| Rutherford score |  |  |  |  |  |  |  |  |
| 4 | 40 | 25.3 | 49 | 28.3 | 50 | 21.3 | .119 | .595 |

|  |  |  |  |  |  |  |
| --- | --- | --- | --- | --- | --- | --- |
| 5 | 114 | 72.2 | 112 | 64.7 | 177 | 75.3 |
| 6 | 4 | 2.5 | 12 | 6.9 | 8 | 3.4 |

BMT – Best medical therapy (anti-platelet/anti-coagulant [if indicated] + lipid lowering agent)
IHD – Ischaemic heart disease
IQR - Interquartile range
TIA – Transient ischaemic attack

Initial treatment strategies and 12-month amputation outcomes are shown in Table 2. No significant differences in initial management strategy were identified. At 12 months, patients within the VaLS cohort were observed to have significantly higher freedom from major amputation (90.5%) compared to both the AP (82.1%, p=.027) and PC (80.0%, p=.005) cohorts (Figure 2). The rate of amputation-free survival was also significantly higher in the VaLS cohorts (72.8%) compared to both AP (54.3%, p<.001) and PC (59.6%, p=.007) groups (Figure 3).

**Table 2.** –Comparison of initial treatment strategy and 12-month outcomes.

|  | VaLS Clinic<br>(VaLS)<br>(n=158 patients) |  | Alternative<br>Pathways<br>(AP)<br>(n=173 patients) |  | Pre-Clinic<br>(PC)<br>(n=235 patients) |  | VaLS<br>vs.<br>AP | VaLS<br>vs.<br>PC |
| --- | --- | --- | --- | --- | --- | --- | --- | --- |
|  | n | % | n | % | n | % | p | p |
| First revascularisation<br>strategy |  |  |  |  |  |  |  |  |
| Endovascular* | 95 | 60.1 | 100 | 57.8 | 143 | 60.9 | .951 | .639 |
| Open | 12 | 7.6 | 15 | 8.7 | 21 | 8.9 |  |  |
| Hybrid | 11 | 7.0 | 11 | 6.4 | 22 | 9.4 |  |  |
| Conservative management | 40 | 25.3 | 47 | 27.2 | 49 | 20.9 |  |  |
| Minor amputation | 13 | 8.2 | 23 | 13.3 | 27 | 11.5 | .139 | .294 |
| Freedom from major<br>amputation (12-months) | 143 | 90.5 | 142 | 82.1 | 188 | 80.0 | **.027** | **.005** |
| Amputation-free survival<br>(12-months) | 115 | 72.8 | 94 | 54.3 | 140 | 59.6 | **<.001** | **.007** |

\*Included covered endovascular repair of the aortic bifurcation (CERAB)

**Figure 2.**
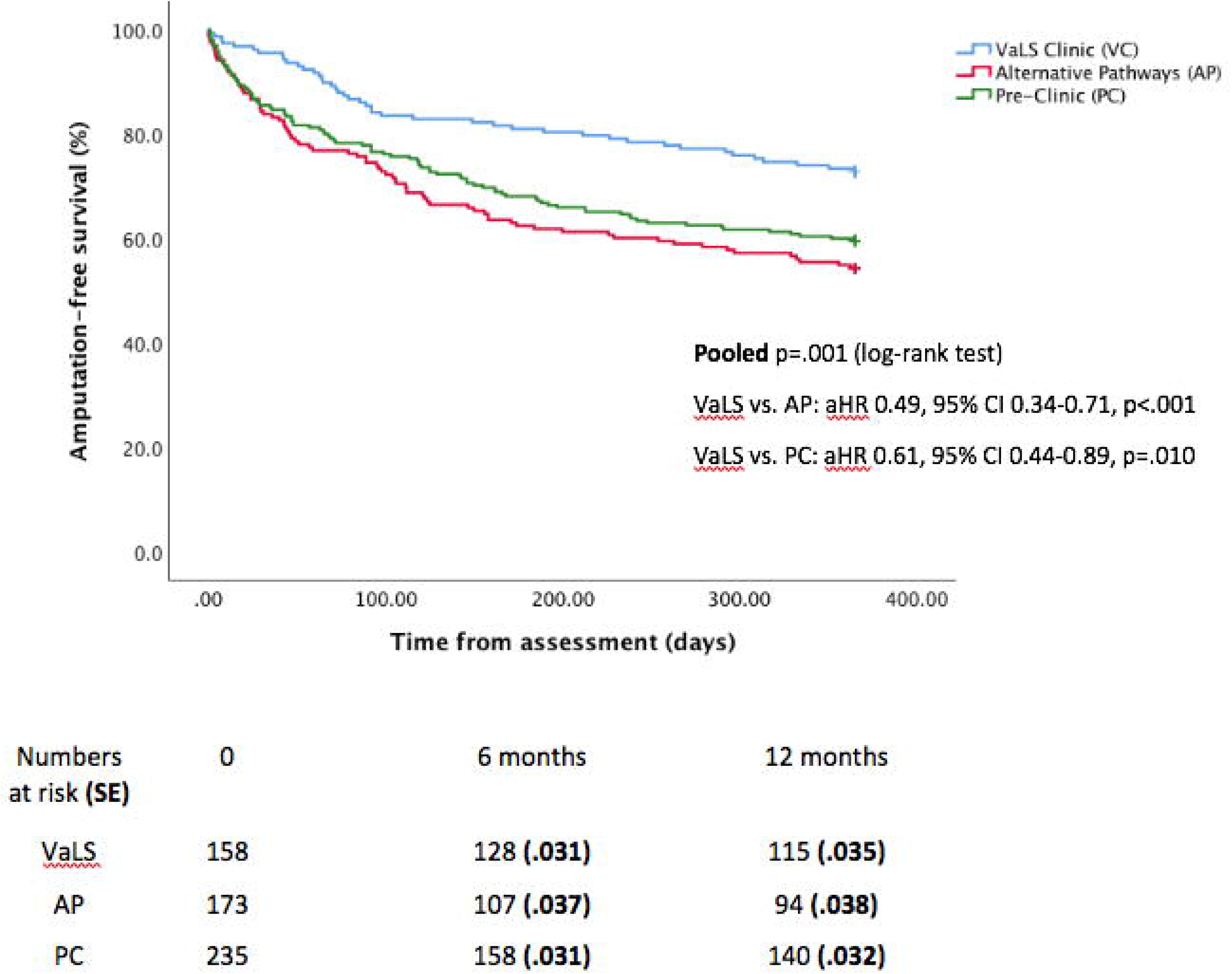
– Comparison of 12-month freedom from major amputation. *VaLS – VaLS cohort, AP – Alternative pathways cohort, PC – Pre-clinic cohort*

Figure 3 – Comparison of 12-month amputation-free survival

*VaLS – VaLS cohort, AP–- Alternative pathways cohort, PC – Pre-clinic cohort*

A final adjusted model was created adding patient factors of age, diabetes, disease severity (Rutherford score), and antiplatelet and lipid lowering therapy use, in addition to patient cohort. Based upon this, the risk of major amputation was significantly lower for patients within the VaLS cohort, compared to both the AP (aHR 0.52, 95% CI 0.28-0.98, p=.041) and PC (aHR 0.50, 95% CI 0.28-0.91, p=.022) cohorts. The combined risk of amputation or death (amputation-free survival) was also significantly lower within the VaLS cohort than both the AP (aHR 0.49, 95% CI 0.34-0.71, p<.001) and PC (aHR 0.61, 95% CI 0.44-0.89, p=.010) groups.

### Timings – assessment to first attempted revascularisation

Following referral, patients within the VaLS cohort were assessed within a median of 2.1 [1.1-3.1] days. For those requiring revascularisation, the median time from assessment to procedure was 6.1 [3.9-11.1] days. Overall, the median time from referral to revascularisation for patients within the VaLS cohort was 8.1 [6.1-14.3] days, with 73.7% (87/118) of patients undergoing revascularisation within the 14-day VSGBI target. For patients with moderate to high risk of amputation (WIfI stage ≥3), 80.9% (55/68) underwent revascularisation within the 14-day target, although only 10.8% (4/37) of those with the highest risk of amputation (WIfI stage 4) met the 5-day referral to revascularisation target.

Upon comparison of median times from assessment to treatment between cohorts, no significant difference between the VaLS, and comparator cohorts, although the spread of times was smallest in the VaLS cohort (Table 3).

**Table 3.**
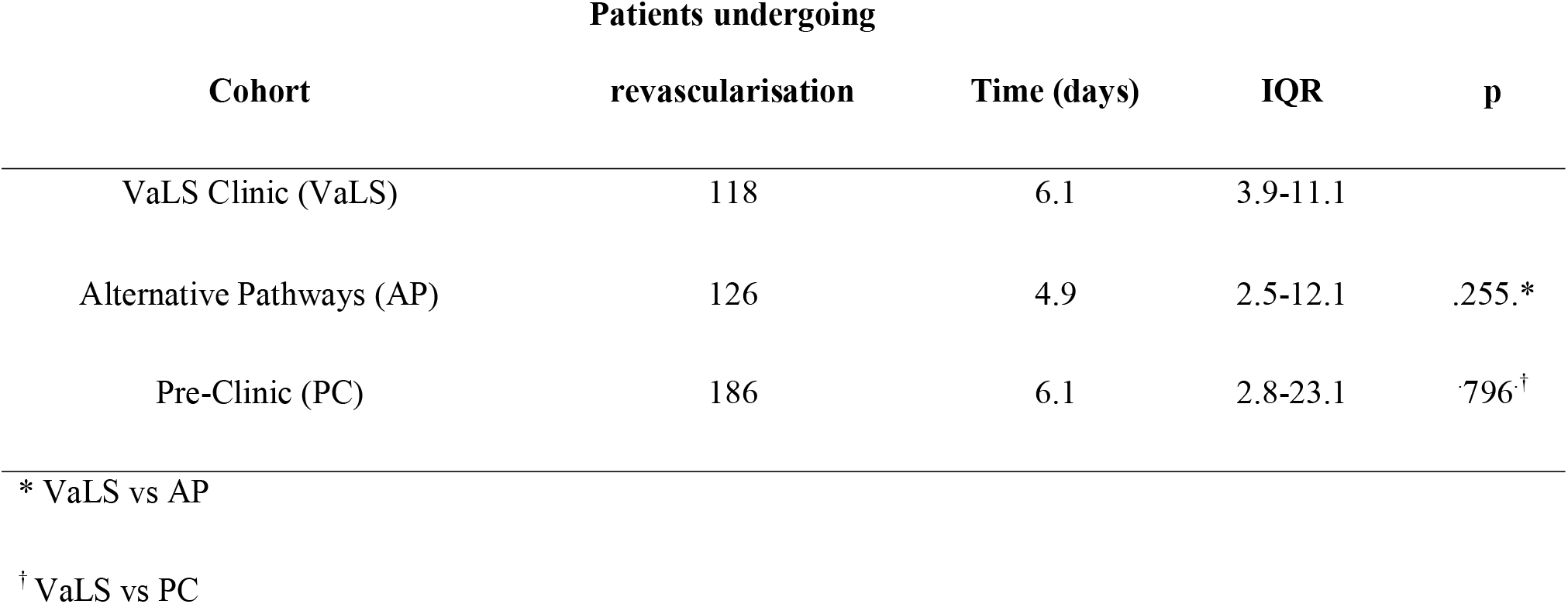
–Comparison of times from assessment to first attempted revascularisation.

## Discussion

This study indicates that the implementation of a novel vascular limb salvage clinic can potentially help to significantly reduce rates of major amputation and improve amputation-free survival in patients with CLTI. Furthermore, these results demonstrate the ability of the clinic to provide timely vascular assessment and consultant review following referral.

How care is provided to this group of patients is an area of growing importance, particularly with the burgeoning prevalence of diabetes mellitus and its complications.^14^ The provision of a vascular service, which facilitates prompt assessment and revascularisation are therefore key aims of any department. Despite this, evidence suggests time delays are common throughout the treatment pathway.^3^ While the causes of delays will differ between individual departments, removing unnecessary barriers between healthcare specialities is appreciated as a key factor in providing timely care.^4, 6^

The concept of vascular limb salvage services has grown in prominence over the last decade as a solution to this problem, providing seamless access to specialist vascular assessment, revascularisation and surgical debridement (often referred to as the ‘toe-and-flow’ model).^8, 9^ The 2019 Global Vascular Guidelines^10^ provides further impetus by defining broad criteria for limb salvage ‘centres of excellence’. Within the contemporary literature, examples of services which meet these criteria are limited and most fail to provide comparative data or adjust for potential confounding (particularly disease severity), thus limiting the validity of any conclusions. Furthermore, individual models vary greatly in scope, design and size; from individual ‘hot’ clinics specialising solely in DFUs, to dedicated multi-aetiology wound-care services.^11^ Within the United Kingdom, focussing on new models of care is highly relevant given the particularly challenging targets recently published within the new VSGBI PAD QIF.^7^

The Leicester VaLS clinic represents one of the first open-access, outpatient based vascular limb salvage clinics which manages patients with CLTI within the literature. Of particular benefit, this model of care provides a readily reproducible and achievable limb salvage service, which utilises many existing resources within a vascular department. The resultant streamlined service, particularly establishing rapid-access to vascular assessment, can potentially help to significantly improve amputation outcomes for patients.

Whilst early results are encouraging, on-going evaluation of the clinic is required to help overcome current and future challenges. Particular focus is necessary to help bring about further improvement in times-to-treatment. Average times from assessment to treatment were comparable between the three cohorts, although lower than those reported within similar studies.^15, 16^ Therefore, whilst the clinic meets the 14 days PAD QIF target, it still falls short of the five-day deadline for patients with the highest risk of amputation (i.e. WIfI stage 4).

This can be interpreted in two ways. Firstly, the ability of the outpatient clinic to achieve comparable treatment times to those managed as inpatient, whilst helping to lower the rate of major amputation is encouraging. This also confers potential financial benefit through reduced use of costly inpatient services. Furthermore, the clinic provides a thorough, outpatient-based vascular assessment and consultant review within just two days of referral.

However, the comparability of treatment times is somewhat surprising given the structure and design of the service. In part, this may reflect current departmental service provision. Currently the clinic does not open at weekends and only emergency revascularisation procedures are performed outside of ‘normal working’ hours. The resultant effect is to potentially delay treatment in some patients upwards of 48 hours, when the time delay is analysed for ‘working days’ time from referral to revascularisation is five days. Of note, accurately identifying the true date of assessment (where a decision to treatment a made) was challenging for the comparator cohorts and therefore these the results may be an underestimation of the true time to treatment.

In light of this, consideration is being given to extending the VaLS clinic to a 7-day service. Furthermore, providing greater capacity for non-emergency weekend operating, as recommended in the recent speciality ‘Getting It Right First Time’ (GIRFT)^17^ report, may help reduce times-to-treatment further. This remains challenging however in the face of ongoing resource constraints and workforce shortages in the National Health Service.^18^

Other results are also noteworthy. Fewer than 50% of patients with CLTI within the VaLS cohort had been prescribed best medical therapy at the time of assessment and the proportion receiving lipid lowering therapy at the assessment were significantly lower in the VaLS cohort. It has long been established that the medical management of PAD, within both primary and secondary care setting, is suboptimal.^19, 20^ These prescription rates are comparable to a recent multi-centred, audit of secondary care practice within the UK and further highlight the need to improve the medical management of patients with CLTI.^21^

Why particularly those within the VaLS cohort are receiving suboptimal medical management is unclear. In part, this may be explained by how prescription data were collected within the clinic (i.e. prospective rather than retrospective collection from electronic patient records) and therefore may be a more accurate assessment of true prescription and compliance rates. Additionally, patients were often referral to the clinic from non-prescribing healthcare professionals, meaning there was less opportunity for them to receive prescriptions prior to assessment in the VaLS clinic.

### Strengths and limitations

In comparison with similar articles reporting outcomes of limb salvage services, this study has two particular strengths. Firstly, it provides comparative institutional data which gives context to the results and helps to demonstrate changes in outcomes. Secondly, the study also provides time-to-event data, adjusts for potential confounding factors and is the first to analyse times within treatment pathways.

Limitations however exist, most notably the potential for selection and performance bias. Whilst baseline characteristics, disease severity and treatment strategies are broadly comparable, CLTI is a complex disease and it was not possible to identify or adjust for all potential confounding factors due to the limitations on the availability data. It therefore stands that patients within the VaLS cohort had an intrinsically lower risk of major amputation. Likewise, decisions on treatments could have been systematically different, with a higher threshold for amputation taken within the clinic compared to the AP and PC cohorts.

In part, this is inherent in any pragmatic analysis of a healthcare service and is difficult to overcome. Understandably, randomised trials in this context would be challenging and unethical and it is difficult to completely remove any performance bias which may benefit those managed within the VaLS clinic. As such, the results of this study should be treated cautiously.

Secondly, the study relied on the accuracy and completeness of HES data and electronic patient records. This was especially important in the context of case ascertainment, identification of patient characteristics and establishing outcomes. Whilst broadly validated, HES data is not without drawbacks including variation in coding between institutions and missing data.^22^ Furthermore, electronic records do not accurately store referral details, clinical examination findings or medication use at the time of assessment, thus preventing the comparison of these factors. Establishing firm assessment dates for patients within the comparator cohorts was also highly challenging retrospectively, meaning the true assessment to revascularisation for these groups may be underestimated. To some extent these effects were moderated by cross checking data with local vascular registry records, however the inability to compare referral times limits the study’s capacity to show definitive improvement since inception of the clinic.

Lastly is the issue of cross-over between cohorts. Whilst not unexpected given the nature of CLTI and the high rate of re-intervention associated with the condition, this cross-over is problematic.^23^ To help manage this, outcomes were assessed on an ‘intention-to-treat’ basis, with patients analysed within the cohort of their first assessment. Therefore, this has the potential of biasing the results in favour of more contemporary patients. The effects of this could have been reduced by selecting comparative cases (namely the PC cohort) from a more distant time-point, however given changes in treatments, this would have affected the comparability of the cohorts.

## Conclusions

The inception of a vascular limb salvage clinic can potentially help to reduce the rate of major amputation for patients with CLTI and provides a model of care which can deliver timely specialist vascular assessment. These findings support the use of vascular limb salvage ‘centres of excellence’ for managing patients with CLTI, as recommended within the Global Vascular Guidelines. Whilst early outcomes are promising, further evaluation is required to assess longer-term outcomes and continue to reduce times-to-treatment.

## Data Availability

Further details on methods, data and materials used will be considered upon email request.

## Acknowledgements

The authors would like to thank George Davies and the George Davies Charitable Trust for the generous charitable donation which funded this work. A pre-print version of this manuscript has been submitted for archiving at www.medrvix.org (doi: 10.1101/19013037v1)

## Previous communication

Presented as an oral presentation at the British Society of Endovascular Therapy (BSET) 2019 Annual Conference (Wotton-under-Edge, United Kingdom, 27^th^-28^th^ June 2019) and awarded the ‘BSET Peripheral Prize’. Presented as an oral presentation in the ‘PAD Scientific Session’ at the 33^rd^ European Society of Vascular Surgery (ESVS) Annual Meeting (Hamburg, Germany, 24^th^-27^th^ September 2019). Presented as an oral presentation at the Vascular Society of Great Britain and Ireland’s (VSGBI) 2019 Annual Scientific Meeting (Manchester, United Kingdom, 27^th^-29^th^ November 2019) and nominated for the Sol Cohen Prize.

## What does this study add?

The introduction of a dedicated vascular limb salvage service within a UK tertiary vascular unit was associated with improved 12-month amputation outcomes for patients with the chronic limb-threatening ischaemia, compared to those managed through traditional clinical pathways. These findings support the use of vascular limb salvage ‘centres of excellence’ for managing patients with CLTI, as recommended within the Global Vascular Guidelines.

## Ethical approval

Approval for this study was granted by the University Hospital of Leicester NHS Trust’s Clinical Audit department (reference 9665).

